# Making Words Count with Computerised Identification of Hypertrophic Cardiomyopathy Patients

**DOI:** 10.1101/2021.04.13.21255353

**Authors:** Luke T Slater, William Bradlow, Trupti Desai, Amir Aziz, Felicity Evison, Simon Ball, Georgios V. Gkoutos

**Affiliations:** Institute of Cancer and Genomic Sciences, College of Medical and Dental Sciences, University of Birmingham; Institute of Cardiovascular Sciences, College of Medical and Dental Sciences, University of Birmingham; University Hospitals Birmingham, NHS Foundation Trust; NIHR Experimental Cancer Medicine Centre; NIHR Surgical Reconstruction and Microbiology Research Centre; NIHR Biomedical Research Centre; MRC Health Data Research UK (HDR UK) Midlands

**Keywords:** text mining, natural language processing, hypertrophic cardiomyopathy, population health management

## Abstract

**Background:** The traditional outpatient model in hypertrophic cardiomyopathy (HCM) is under pressure. Population health management based on an accurate patient record provides an efficient, cost-effective alternative.

**Methods:** To improve the accuracy of the HCM patient list in a single hospital, we developed a rule-based information extraction natural language processing (NLP) framework. The framework employed ontological expansion of vocabulary and exclusion-first annotation, and received training by an ‘expert in the loop’. The output stratified patients with atrial fibrillation (AF) and heart failure (HF), those without active cardiology care and likely screened individuals.

**Results:** The algorithm was validated against multiple data sources, including manual validation, for HCM, AF and HF and family history of the disease. Overall precision and recall were 0.854 and 0.865 respectively. The pipeline found 25,356 documents featuring HCM-related terms belonging to 11,083 patients. Excluding scanned documents resulted in 17,178 letters from 3,120 patients. Subsequent categorisation identified 1,753 real cases, of whom 357 had AF and 205 had HF. There were 696 likely screened individuals. Adjusting for 304 false-negative patients, the total HCM cohort was 2,045 patients. 214 were not under a cardiologist. NLP uncovered 709 patients who were absent in the registry or hospital disease codes.

**Conclusion:** This novel NLP framework generated a hospital-wide record of patients with HCM and defined various cohorts, including the small set of HCM patients lacking current cardiology input. Existing data sources inadequately described this population, spotlighting NLP’s essential role for clinical teams planning to move to a population health management model of care.

## Introduction

Hypertrophic cardiomyopathy (HCM) affects between 0.2%^1^ and 1.4 %^2^ of individuals and is an important cause of sudden death, atrial fibrillation (AF) and heart failure (HF). Patient numbers are growing, driven by an expansion in those with mild disease^3^. The fixed one to two yearly outpatient appointment model ^4,5^ was strained even before the COVID-19 pandemic. Population health management (PHM) offers a solution to these challenges ^6^ but relies on an accurate record of diagnosed individuals.

Clinical teams can find HCM patients within automatically generated clinic lists, but this is time-consuming. The most accessible record comes from routine hospital statistics, but this contains only patients with prior inpatient episodes. Disease registries can be repurposed but are prone to missing patients. Developing, populating and maintaining them is also laboursome and error-prone. Searching the electronic record by hand using keyword terms is the most arduous option.

Natural language processing (NLP), which describes the automated understanding and generation of human language, is a field of research in computer science relevant to population health management^7^. While applications in cardiovascular disease are spreading^8–11^, publications in HCM are limited^12,13^.

Our study’s aim was to improve the accuracy of a single hospital’s list of patients with HCM. To do so, we set out to:

- expand our existing NLP algorithm^13^, deploy it across the electronic patient record and test its accuracy
- define the entire cohort, subgroup by AF and HF complications, identify which patients needed to see a cardiologist and find people likely being screened
- measure the impact NLP had on total headcount compared to existing data sources

## Methods

We undertook this work at the Queen Elizabeth Hospital site of University Hospitals Birmingham NHS Foundation Trust. The authors declare that all supporting data are available within the article (and its online supplementary files).

### Information Extraction Framework

Figure 1 depicts the seven steps that make up the information extraction pipeline. We implemented the pipeline using the Komenti semantic text-mining framework^14^, a natural language processing program developed in-house. This is available under an open-source licence at https://github.com/reality/Komenti.

**Figure 1.**
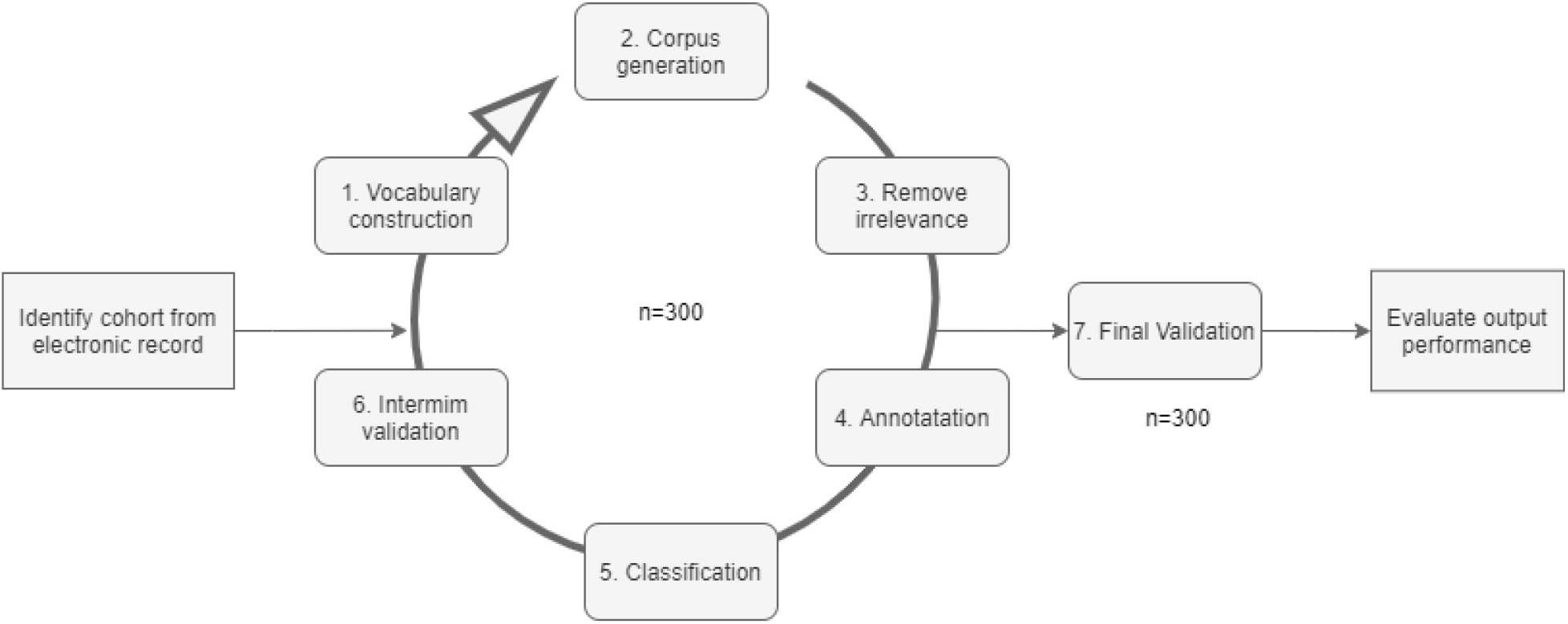
Natural Language Processing Pipeline for Identification of Patients with Hypertrophic Cardiomyopathy. The testing cycle was repeated until high performance was achieved

#### 1. Vocabulary Creation

We first developed a vocabulary of diagnoses and their synonyms that identify the conditions of interest in the text. A clinical specialist (WB) produced a list of words and phrases that indicate HCM, AF and HF (see Appendix for the full list). The vocabulary contained phrases that pointed to a disease complication, for example ‘weak heart’ for HF. Komenti’s vocabulary expansion algorithm broadened the number of searched-for labels and synonyms through linkage to biomedical ontologies.

#### 2. Corpus Generation

Documents were obtained from the UHB patient document management system (OpenText) on 02/06/2019. The entire clinical document record, around 22TB in size, was collected on a server in PDF format and queried using a keyword search to identify documents that included labels for HCM. The corpus included referrals from primary, secondary and tertiary care, clinic letters, discharge summaries and typed digital noting. Cardiac imaging reports were excluded since they are stored outside OpenText.

A minority of these documents were scanned. This included all Emergency Department documents and genetic reports. Examination of scanned documents used an optical character recognition extraction tool found within the OpenText suite. Assessment for complications only included the most recent document mentioning HCM together with AF and HF.

#### 3. Removal of irrelevant data, initial term recognition, and tokenisation

After identifying documents matching HCM keywords, we ultimately removed scanned documents which were a source of numerous errors (described in Results; Interim Validation). We then tokenised the remaining documents into sentences. Terms from the vocabulary pertaining to HCM, AF, and HF were found in the sentences, and only sentences containing those keywords were retained. We then developed a vocabulary for the exclusion of sentences that contained irrelevant mentions of HCM, AF, and HF; in which the text did not actually refer to the condition of interest (See Results; Interim Validation for examples). This step also involved excluding acronyms that could be confused for other conditions.

#### 4. Annotation

The pipeline grouped sentences by scenarios where mentions did not indicate definite disease for the patient, before eliminating them to leave only genuine cases. The three groups were:

- Negated: The patient does not have the condition
- Uncertain: The patient might have the condition
- Family history: The sentence refers to a family member and not the patient

The negation and uncertainty detection algorithms employ a graph-based dependency extraction method to quantify the grammatical proximity between the tag and the word of interest^13^. A bespoke vocabulary of negation words was developed with the clinical specialist. The uncertainty instance was based on a resource originally intended for topic modelling^15^ and improved alongside the clinical specialist.

The dependency-based parsing approach disambiguated complex individual sentences featuring more than one condition of interest (Figure 2). For example, in the sentence “*the patient has HCM, but does not have atrial fibrillation”*, the algorithm would link ‘*not’* to ‘*AF’*, rather than ‘*HCM’*.

**Figure 2.**
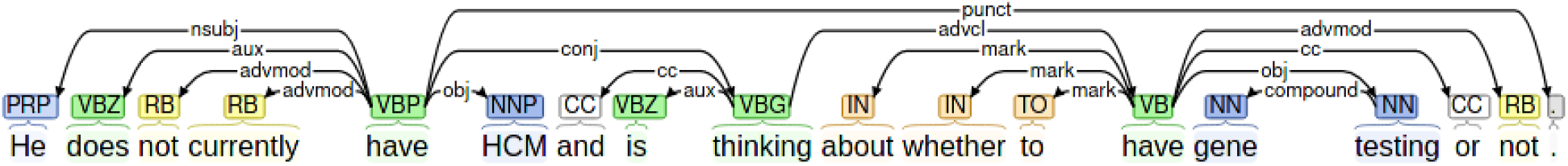
Dependency Parsing Tool Deconstructs Hypertrophic Cardiomyopathy (HCM) Mention Containing Sentence. The tool determines the strength of relationships between modifiers and mentions to arrive at the context in which the mention is made.

To identify mentions where there was a family history of the disease, we took a simple approach where the mention of family vocabulary words in the sentence (brother, sister, father, mother etc) was considered sufficient to mark the sentence as referring to a family member. After matching each sentence to a context, it was annotated with a tag.

#### 5. Classification

The framework determined the patient’s final status for each condition by adding context indicators to sentences. Where there were multiple sentences expressing conflicting evidence about the condition, the final decision was arbitrated by counting which context occurred most frequently (see Figure 3). The framework repeated the process for family relations mentioned in letters.

**Figure 3.**
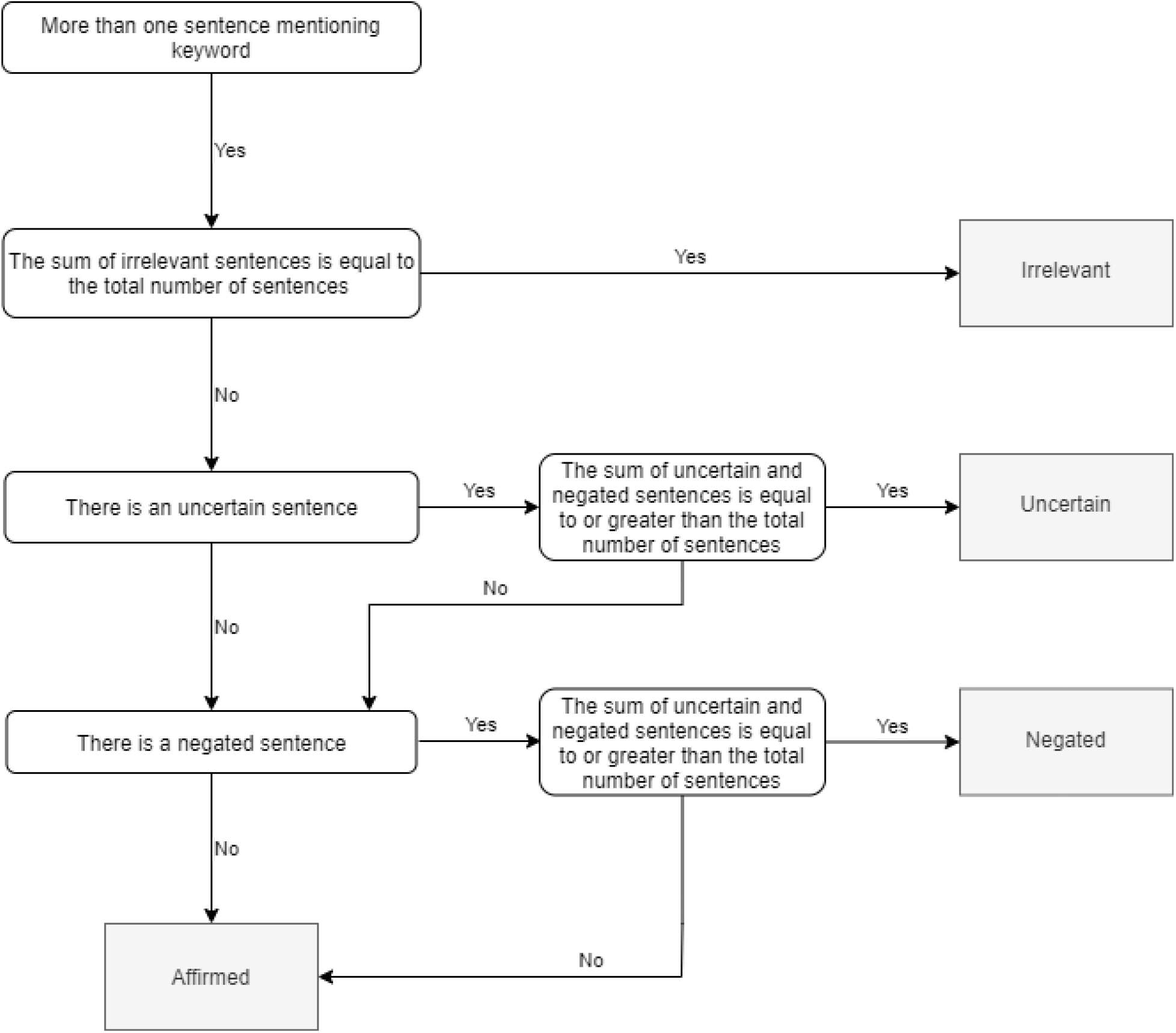
Flow Diagram to Resolve Conflicting Evidence at Classification Stage.

#### 6. Training with Interim Validation

A simple web server and client tool was developed for clinicians to validate the results of the algorithm-generated annotations. The tool surfaced all relevant documents for a set of patients and condensed them into a single-page web application (Figure 4). This displayed the algorithm’s judgment for HCM and comorbidities alongside the evidence for the decision, based on an excerpt from the clinic letter. Space was provided for free text feedback at the bottom of the page.

**Figure 4.**
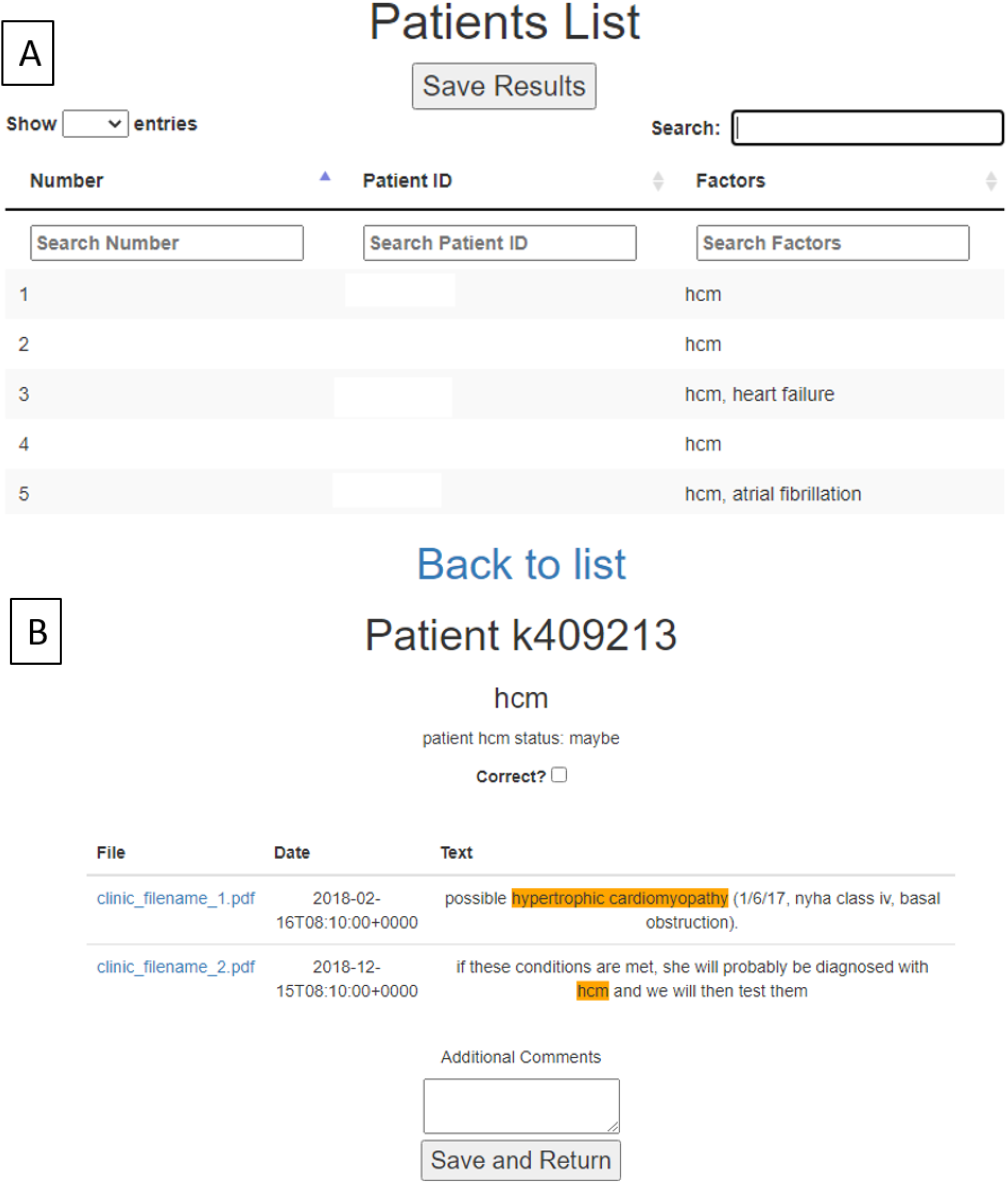
Web Server Validation Tool for Hypertrophic Cardiomyopathy (HCM) Patients. A, Summary page displaying list of patients with HCM-related documents itemised by search number and patient ID and filterable by search factors; B, Patient level page showing algorithm’s judgment, evidence for the decision, box for users to check whether the algorithm performed accurately or not, option to view the pdf of the document in question and space to collect free text feedback. Figure shows data synthesised for demonstration, not pertaining to any patient.

The algorithm’s performance was evaluated on a patient by patient basis and results saved as json files for export. For each experiment, training on 300 patients took place over several iterations of the pipeline using feedback from a clinical specialist (WB).

The interim validation results determined whether the algorithm had correctly assigned the status (uncertain, negative, family history, positive) for each of the concepts based on the text provided. It also allowed feedback for cases where algorithm classification had been correct, but the vocabulary has been incorrect or non-specific. These results informed modifications to each of Steps 1 - 5, particularly vocabulary and classification. Repetition of interim validation occurred until the training set achieved high precision and recall.

#### 7. Algorithm Evaluation by Internal Validation

We validated the pipeline outputs in a series of experiments against three structured sources; a clinico-genomic registry used to collect data on HCM patients in the Heart Muscle Disease Service since 2015, International Classification of Diseases,Tenth Revision (ICD-10) codes from the Hospital Episodes Statistics (I421 and I422) and a database of automated ECG reports. The first two contained information on HCM, AF and HF while the last resource related to AF only.

When validating HCM comorbidities, we only used records for individuals who had both HCM and disease complications featured in ICD-10 codes and the registry. The definition of HF in the registry was evidence of current heart failure or a prior admission with heart failure. Interrogation of the automated ECG reports and ICD-10 codes took place before extraction of the corpus. Consequently, validation against these databases excluded patients whose clinical documents in the corpus extraction were created after these cut-off dates.

The clinical specialist (WB) further validated an additional 300 patients excluded from the training set by hand.

Performance was measured using precision (true positives/true positives + false positives) and recall (true positives/true positive + false negative). Precision derived from manual validation, and recall from validation against databases. We excluded from experiments several instances where the validation would be flawed and produce misleading performance values:

- precision measurements versus databases, as the databases do not cover every patient (i.e. are not gold-standard comparators).
- recall measurements versus manual validation, as their inclusion would distort how well the algorithm performed at finding the true number of cases.
- recall measurements versus databases in patients that Komenti did not classify, i.e. patients that the document search did not find sentences mentioning HCM. This situation arose when the diagnosis lay outside the document corpus, or in an unusable form within the corpus (i.e. scanned documents, see Result, Interim Validation).

It took two days to collect the corpus and an hour for the pipeline to run.

### Population Definition and Stratification

We identified the entire cohort, stratified it and found which patients needed to see a cardiologist using the following stages:

Stage 1. Removed scanned documents from the total number of documents with HCM keyword mentions.

Stage 2. Confirmed cases identified by the algorithm after filtering irrelevant, negative or uncertain mentions, before subcategorizing for AF and HF. Patients who were negative or uncertain for HCM but had a family history of the condition were assumed to have been screened.

Stage 3. False negatives (HCM patients present in the registry and ICD-10 groups which NLP had not found) were added to the NLP-found group to produce the total HCM cohort.

Stage 4. Surviving HCM patients who were not under active cardiology follow-up were found by dividing the total HCM cohort into the following groups using administrative data:

- Dead
- Lost to follow up (patient did not return for continued care as expected, defined here as seen in the outpatient setting over 24 months ago with follow up arranged but not seen since, not on a waiting list, not discharged)
- Under active care (under a specialist cardiologist in the Heart Muscle Disease or Inherited Cardiac Conditions Service, a general cardiologist or a non-cardiologist)
- Seen as an inpatient only
- Discharged (and not under active care in another service).

Patients on a follow-up waiting list were defined as being currently under active care. New patients awaiting their first consultation were not included. We defined patients who need to see a cardiologist as those lost to follow up, those under active management who were not under cardiology or those seen as an inpatient only.

### Comparison of Data Sources

The total cohort was divided into three groups to determine the contribution made by NLP; Group 1 (registry), Group 2 (ICD-10, not in Group 1), Group 3 NLP (NLP found, not in group 1 or 2).

### Ethics

The University of Birmingham Ethical Review granted approval (ERN_20-0338). No informed consent was required as this was a service improvement project, and the documents were not de-identified as we intend to follow up individuals lost to follow up. The hospital number was used to link a patient’s clinical documents to their registry record. Data remained within the Trust and only included information related to HCM.

## Results

### Interim Validation

The training phase revealed various errors (Table 1); the commonest arising in scanned documents where OCR misread a fragment of a word as an entity of interest (Figure 5). Consequently, we excluded all scanned documents, including those from the Emergency Department and the West Midlands Regional Genetics Service. Irrelevant documents were also cast out, for example when the document header contained the entity of interest; “*Heart Failure* clinic” (Figure 5). To address this problem, we adjusted the pipeline to detect addresses, phone numbers and postcodes.

**Table 1.** Key Issues Identified During Training on Hypertrophic Cardiomyopathy (HCM) Keyword-Containing Documents with Response.

| Error | Example | Response |
| --- | --- | --- |
| Misidentification of distorted text | HCM identified within garbled sentence in scanned PDF | Removal of scanned documents from corpus |
| Irrelevant sentences found in header and footer | ‘Heart failure’ identified as part of the address of a clinic, named ‘heart failure clinic’, or the abbreviation HCM identified within the hospital’s chief executive name | Exclude sentences that contain phone numbers, or postcodes |
| Extraneous vocabulary | ‘diastolic dysfunction’ is an unreliable indicator for heart failure as it captures individuals with only mild diastolic dysfunction but no heart failure | Refine vocabulary |
| Missing vocabulary | ‘Possible HCM’ is not marked as uncertain, because *possible* is not in the uncertainty vocabulary | Extend vocabulary |
| Algorithm failure | Algorithm fails to identify a negated concept | Improvement of the algorithm |
| Text pre-processing | Text parsed from PDFs missed whitespace and combined several words into one | Moved to a different library for parsing PDFs |

PDF indicates portable document format

**Figure 5.**
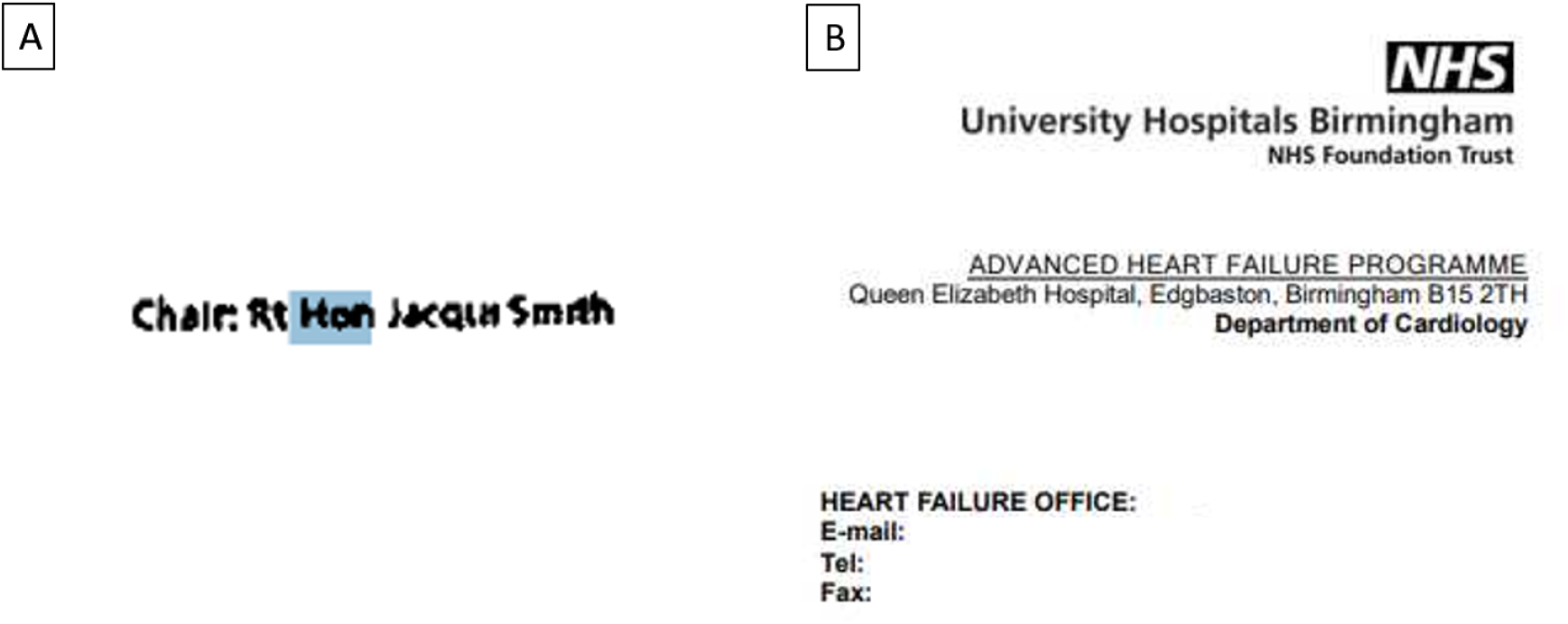
Examples of Irrelevant Mentions Encountered During Pipeline Development. A, Artefact (highlighted) within the Trust Chair’s title which was misread for HCM (hypertrophic cardiomyopathy), created by optical character recognition of a scanned document; B, letter header containing the token Heart Failure.

We had developed the vocabulary to maximise coverage of diagnosed patients but this resulted in a high false-positive rate from non-specific phrases. For example, the algorithm initially flagged Grade 1 diastolic dysfunction as being indicative of heart failure. The vocabulary was updated by excluding these phrases.

### Performance

Table 2 shows the framework’s overall performance measured against three different structured data sources, along with summary statistics for the manual validation.

**Table 2.**
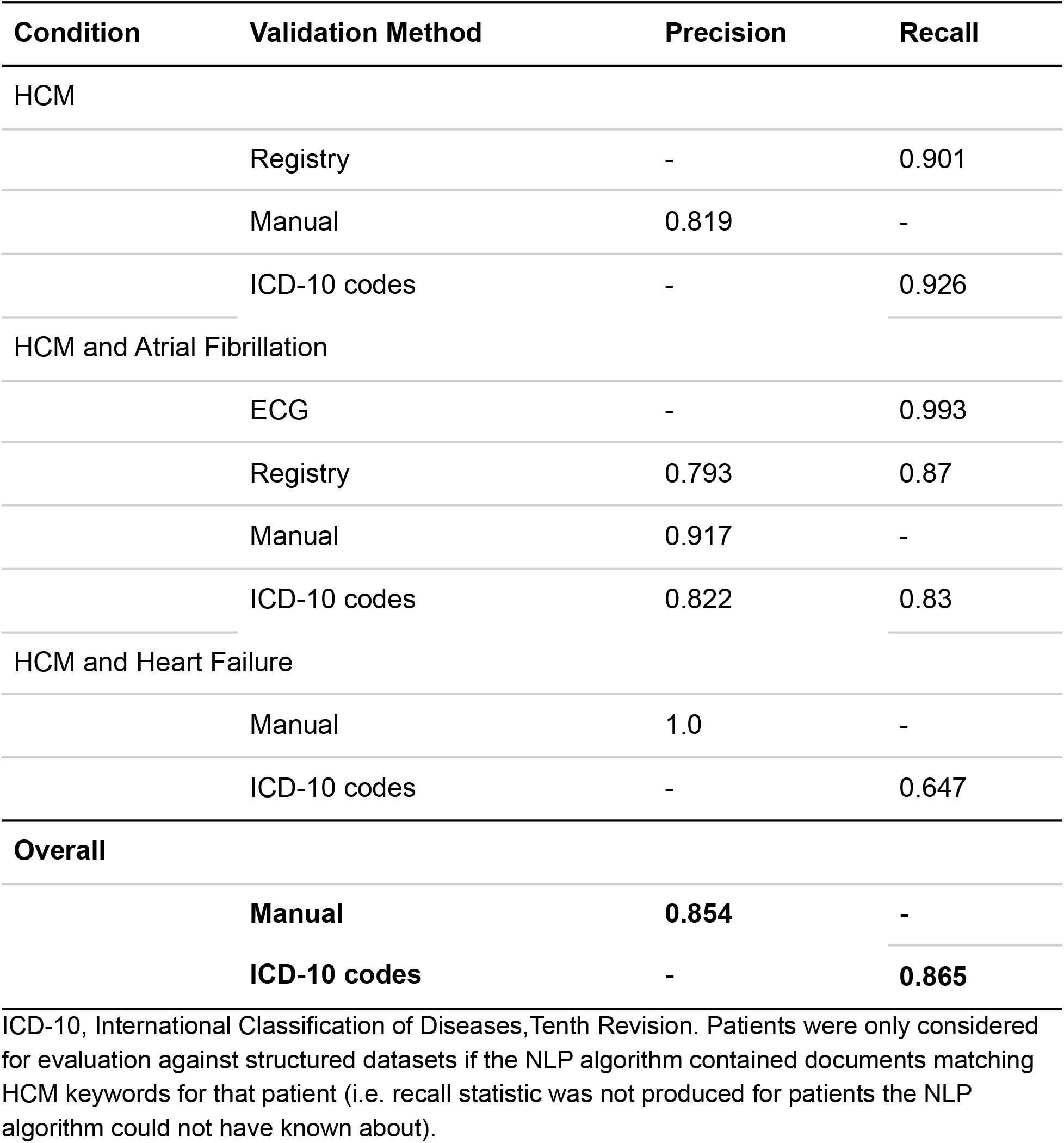
Natural Language Processing (NLP)’s Performance Measured Against Structured Resources in Hypertrophic Cardiomyopathy (HCM) Keyword-Containing Documents.

For the identification of HCM patients, precision and recall were high. Manual validation showed that a small number of negative and uncertain cases were incorrectly classified by the algorithm. For the case of HCM patients with AF, the performance was good in all experiments while for HCM patients with HF recall was lower when validated against ICD-10 codes.

There are two possible explanations for the latter. The first is that clinical coders identified patients with HCM and HF within ICD-10 codes from Emergency Department documentation, or in handwritten inpatient notes, which could not be examined via NLP.

The second is that clinical coders picked up on HF recorded in documents which did not also mention HCM.

Table 3 displays a breakdown of the manual validation performance per classification and shows that good precision persists across the various subcohorts indicating that the framework can accurately stratify patients into HCM-related subgroups. The low precision for patients with HCM and HF relates to the low patient numbers.

**Table 3.**
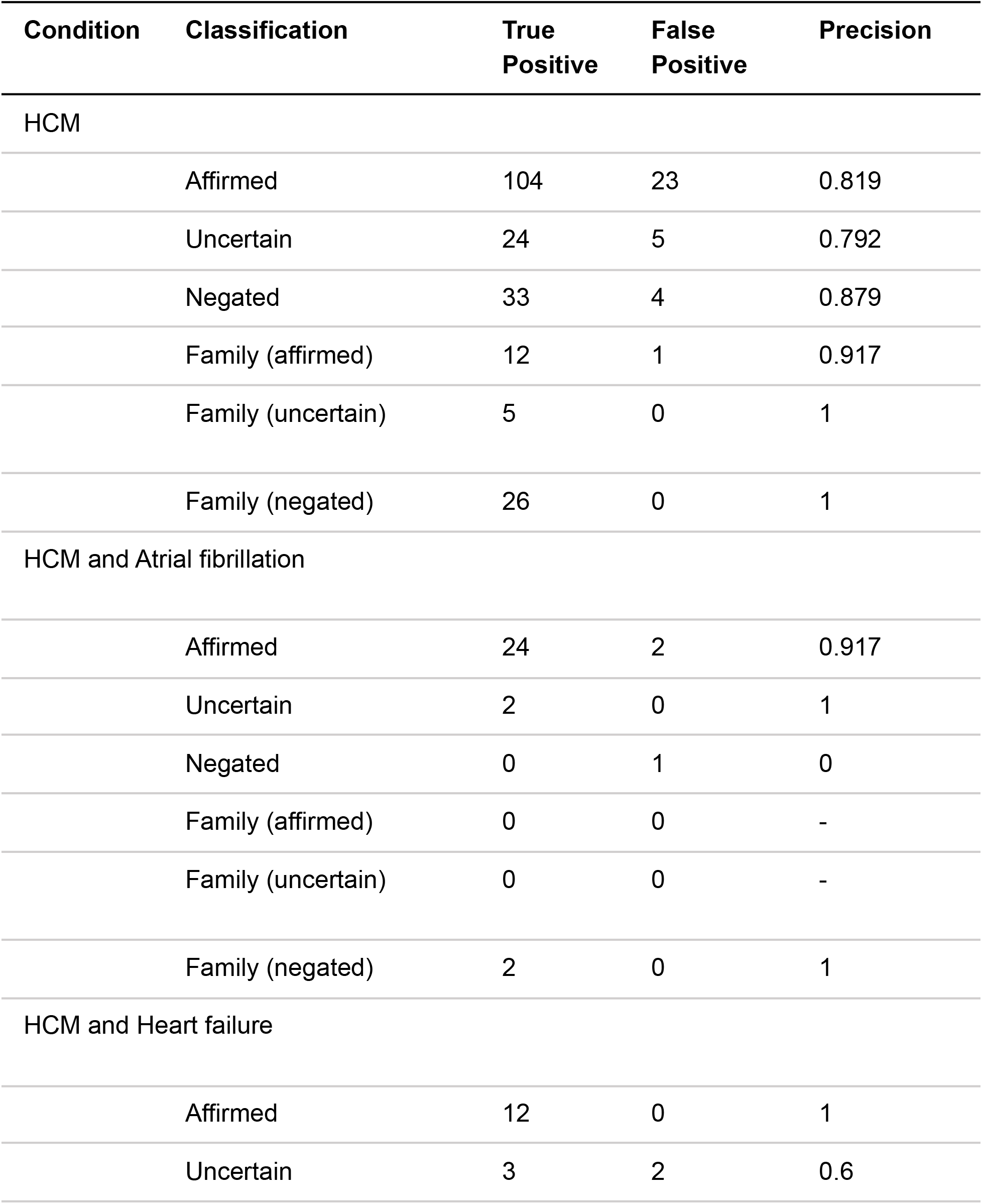
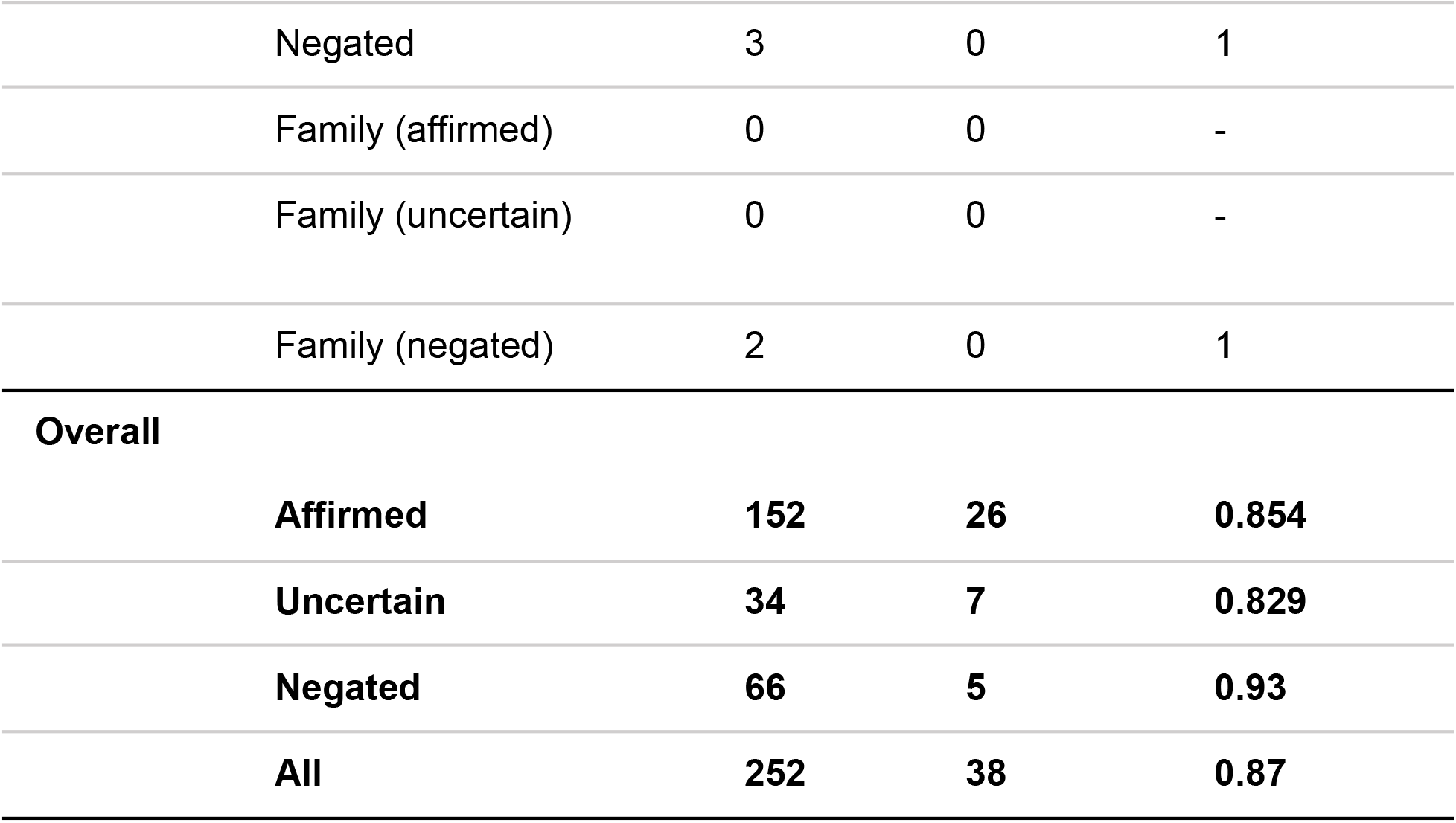
Natural Language Processing (NLP)’s Performance Measured Against Structured Resources in Hypertrophic Cardiomyopathy (HCM) Keyword-Containing Documents by Context.

NLP also improved the quality of registry data by identifying 24 people in whom staff had misentered patients as having HCM in the registry when in fact they were gene-positive but phenotype negative.

### Population Definition and Stratification

In Stage 1, as shown in Figure 6, from a total of 25,356 letters with HCM keywords related to 11,083 patients, 8,178 scanned documents were removed. This left 17,178 documents belonging to 3,120 patients.

**Figure 6.**
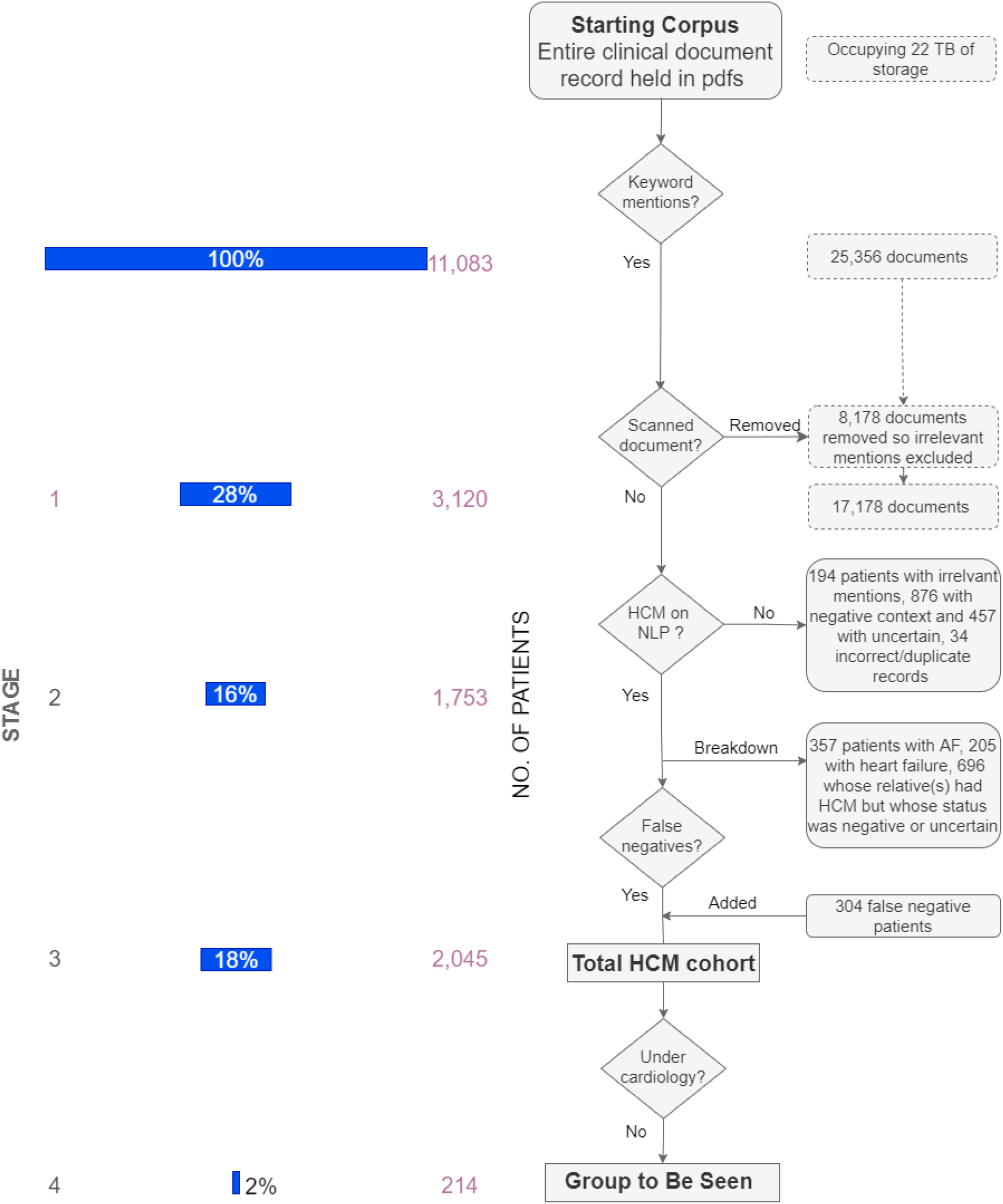
Text-Mining Pipeline Distilling 25,356 Hypertrophic Cardiomyopathy (HCM) Keyword-Containing Documents into a Cohort without Cardiology Care. Patient numbers are provided on the left for each Stage of the pipeline. The boxes with dotted outlines describe document numbers. Boldface indicates key points in the flowchart; NLP, natural language processing.

In Stage 2, the algorithm then classified and excluded 876 patients who were negative for the condition and 457 patients with uncertain disease (Table 4). A further 34 patients were found by the NLP algorithm but then discovered not to exist due to either incorrect metadata in the document system, or incorrect extraction of hospital numbers from letters. They were excluded from further analysis.

**Table 4.** Summary of Patient Classifications in Hypertrophic Cardiomyopathy (HCM) Keyword-Containing Documents.

| **HCM Classification** | **Number of Patients** |
| --- | --- |
| HCM Keywords | 3120 |
| Without HCM (negated) | 876 |
| Uncertain HCM status | 457 |
| Incorrect/duplicated hospital numbers | 34 |
| HCM (affirmed) | 1753 |
| HCM and Atrial fibrillation | 357 |
| HCM and Heart failure | 205 |
| Family History of HCM | 1029 |
| and HCM themselves | 333 |
| and either uncertain or negative status themselves | 696 |

Of confirmed HCM patients, 357 had AF and 205 HF. 1,029 patients were found to have a family history of HCM, 696 of which had an uncertain or negative status themselves.

In Stage 3, 1,753 patients were classified as positive for HCM. 304 patients (91 in registry, 213 in ICD-10 codes) which weren’t found by NLP were added, producing a total cohort of 2,045 HCM patients.

The registry false-negatives arose because our organization has historically provided local hospitals with cardiovascular magnetic resonance (CMR) imaging, and genetic testing via outreach from the neighbouring West Midlands Regional Genetics Service. Although these patients were captured in the registry, they did not have NLP-accessible documents. For ICD-10 false negatives, it is likely that inpatient stays were coded for HCM but excluded from NLP analysis as the diagnostic data was recorded in handwritten inpatient notes and omitted from electronic discharge summaries.

In Stage 4, this information was integrated with administrative data (Table 5). Almost twelve hundred patients were under active management, 361 were discharged and 310 had died. In total, 214 patients were not under active care with a cardiologist.

**Table 5.** Total Hypertrophic Cardiomyopathy (HCM) Cohort Breakdown by Status; with Resulting Number to be Seen.

| **Status** | **No. of Patients (%)** |
| --- | --- |
| Dead* | 310 (15) |
| Lost to Follow Up |  |
| From Heart Muscle Disease Service | 61 (3) |
| From Inherited Cardiac Conditions Service | 30 (1) |
| From General Cardiology | 68 (3) |
| Total Lost to Follow Up* | 92 (4) |
| Under Active Management |  |
| Heart Muscle Disease Service | 981 (48) |
| Inherited Cardiac Conditions Service | 13 (1) |
| General Cardiology | 471 (23) |
| Only by non-cardiologist* | 73 (4) |
| (Total under active management (cardiology))* | 1160 (57) |
| Seen as inpatient only* | 49 (2) |
| Discharged from |  |
| Heart Muscle Disease Service | 145 (7) |
| Inherited Cardiac Conditions Service | 98 (5) |
| General Cardiology | 536 (26) |
| Total Discharged* | 361 (18) |
| **(Total to be seen)** | **(214, 10)** |
| **Total** | **2,045** |

The rows with a (\*) in the final column are mutually exclusive. As the same patient could be seen by multiple teams, the numbers within the team row relate to the patient's current status with that team only, i.e. a patient may be actively monitored by general cardiology but has been discharged from Heart Muscle Disease Service. The total rows for patients not being actively monitored by any cardiology team are based on their last interaction. The total to be seen (bold) is the sum of lost to follow up, seen only by a non-cardiologist and seen as inpatient only.

### Comparison of Data Sources

Subdivision of the confirmed HCM cohort by data source (Figure 7, Table 6) showed that ICD-10 added 404 patients (Group 2) to Group 1, which contained 932 patients. NLP contributed 709 patients (Group 3). The relatively modest provision by ICD-10 codes reflects the fact that coding is only applied to inpatient and not outpatient visits. NLP recapitulated almost all patients captured by the registry and ICD-10 codes, apart from the 304 false negative patients.

**Table 6.** Breakdown of Hypertrophic Cardiomyopathy (HCM) Patients per Data Source.

| **Group/Data Source** | **Number of Patients** |
| --- | --- |
| Group 1 ( In registry) | 932 |
| Found in ICD-10 codes | 777 |
| Found in ICD-10 codes, also in registry | 373 |
| Group 2 ( not in Group 1 but found in ICD-10 codes) | 404 |
| Found Using NLP | 1,753 |
| Found using NLP, also in registry | 480 |
| Found using NLP, also in ICD-10 codes | 203 |
| Found using NLP, also in both registry and ICD codes | 361 |
| Group 3 (Unknown to registry and ICD codes) | 709 |
| Not found by NLP but present in registry | 91 |
| Not found by NLP but present in ICD-10 codes | 213 |
| **Total unique HCM patients (Group 1 + Group 2 + Group 3)** | **2045** |

NLP indicates natural language processing; ICD-10, International Classification of Diseases, Tenth Revision.

**Figure 7.**
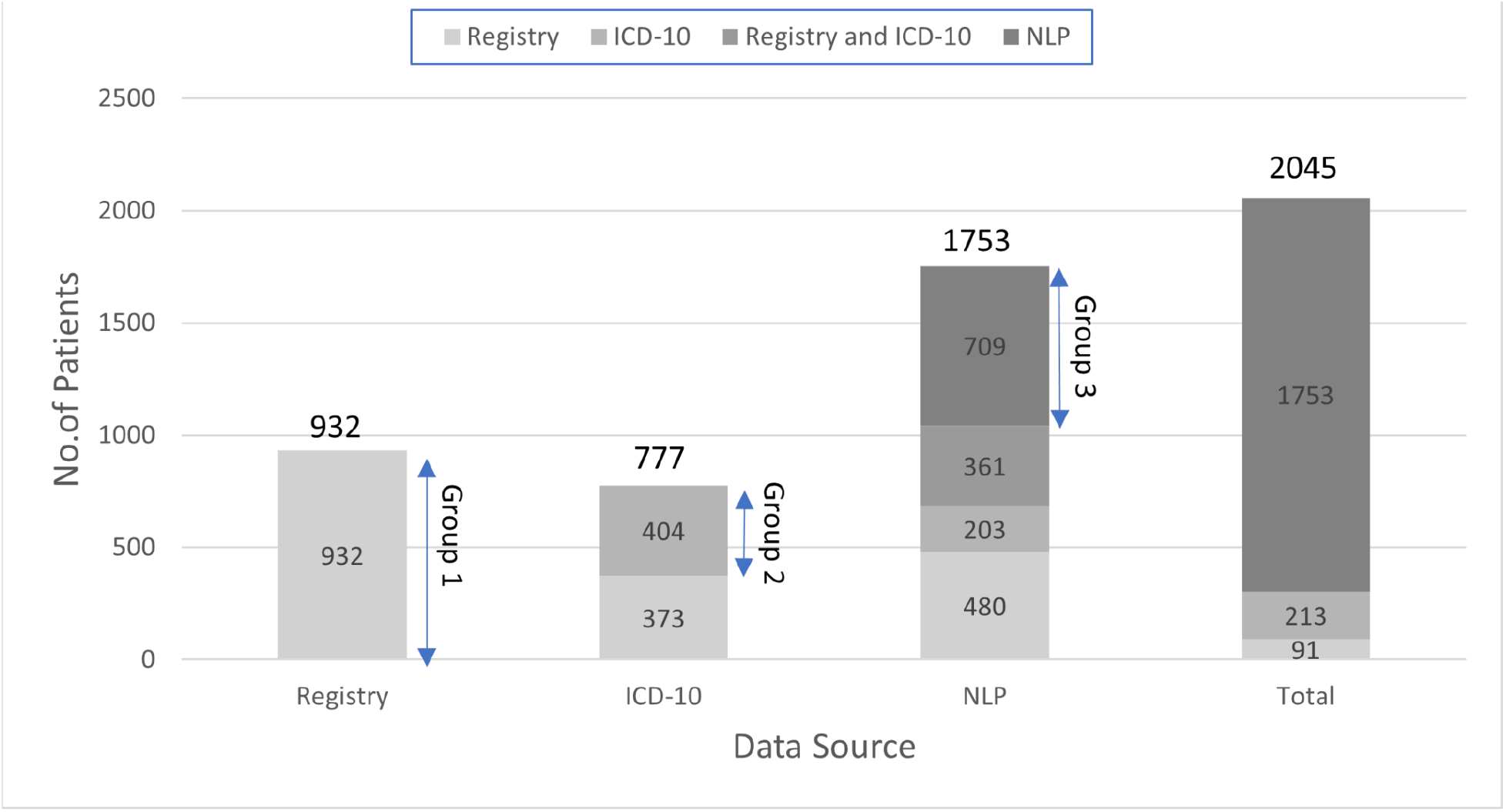
Distribution of Hypertrophic Cardiomyopathy Patients by Data Source. Totals for each data source are given above the bar. Groups 1 (registry), 2 (International Classification of Diseases,Tenth Revision [ICD-10], not Group 1) and 3 (natural language processing [NLP] found, not in Group 1 or 2) are identified by double-headed arrows.

## Discussion

### Need for Population Health Management Model of Care

Specialist HCM services are grappling with three factors. Firstly, demand is rising, with a tenfold increase in the number of diagnoses after 2010 compared to pre-2000^3^. There is an associated shift in demographics and disease features. The average patient is older, less severely affected and lower-risk^3^, likely a consequence of increased disease awareness, imaging and genetic-led screening.

Secondly, the COVID pandemic has uprooted the traditional, fixed-appointment follow-up schedule. Although this model doesn’t prevent patients from being lost to follow-up, it did at least provide a safety net for long-term monitored patients. Finally, while clinical teams have the knowledge needed to re-design services ^16–20^, current electronic records make it difficult to act on this data systematically. Instead, new knowledge is implemented by hand on a patient-by-patient basis.

Population health management offers a means to respond by automatically capturing, stratifying and tracking an entire population of patients to address inequities, intensify care of the sickest and manage the healthiest remotely. Our study examines how NLP might help clinical teams adopt this approach by first generating an accurate and partially-stratified patient list in a large teaching hospital.

### Comparison with Existing Models

NLP frameworks employ either machine learning or classifier recognition/classification strategies. Although machine learning approaches return results quickly after a training set is assembled, we chose the alternative as its basis in the grammatical roots of language makes it disease-agnostic.

Incorporating an expert clinician’s background knowledge to generate vocabulary and rules for concept recognition in text has been shown to out-perform machine learning methods in certain settings^21^. The web validation tool and workflow, in combination with the utilisation of domain vocabularies, enables quick and continuous expert feedback leading to improved performance. For the clinician, the web validation tool markedly shortened the review process. It also allowed them to understand how errors came about as evidence regarding the assertion was presented.

The strategy of determining clear-cut cases by excluding patients whose disease mentions did not imply that they had the condition is typical of negation classifiers in information retrieval pipelines (for examples see NegEx and negation-detection^22,23^). Our approach differs from these methods as it uses a dependency parser distance procedure and includes additional classifiers for sentence context. In a recent head to head comparison with these algorithms, which involved data concerning 5000 sentences from HCM patients at our hospital, ours performed favourably^13^.

The only other work applying NLP in HCM^12^ involved an open-source NLP package called MedTagger. It focused on mining for instances of syncope and positive family history for sudden death within 200 patients with HCM selected from a specialist service registry. Finding diagnosed HCM patients across the entire electronic patient record is a more complicated task than searching within known patients, as evidenced by a lower performance for identification of HCM compared to detection for the two comorbidities.

The Mayo work used a neural network-based approach to information extraction, while our method employs a rule-based system. In specific settings, rule-based NLP systems offer better performance over machine learning-based strategies as they can be tailored to the needs of an experiment^21^. Different validation sources (billing codes for previous work, ICD-10 for ours) also make the results of comparison difficult. In our setting, we found that ICD-10 codes were an inadequate resource for diagnoses.

### Implications for Clinical Services Transitioning to Population Health Management

After casting out scanned documents, NLP was able to place a full list of patients in the clinical team’s sights quickly, minimising the manual work needed to obtain this information. To ensure HCM patients are not missed, the notes of the 1333 negative or uncertain HCM patients could be rapidly reviewed via the web tool. Without this pipeline, evaluating 3,120 patient records featuring over 17,000 HCM-keywords containing documents by hand would be impractical.

Subdividing HCM patients for the two commonest complications means patients can be cohorted in care pathways. We did not attempt the more complex task of stratifying patients by sudden death risk using text mined risk factors ^18,19^. Merging NLP output with follow-up arrangements identified the 1.9% of patients who were at high risk of being entirely overlooked as they were not under a cardiologist.

By determining the small number of particularly vulnerable patients who would benefit from further evaluation, the clinical team could allay concerns regarding the amount of additional work NLP would generate. It’s likely that the final number would be smaller still after adjusting for patients under active cardiology management elsewhere. Care could also be streamlined for likely screened patients, identified as a byproduct of the use of family history in patient annotation.

NLP was able to find almost all of the HCM patient cohort already known to the hospital, highlighting ICD-10 codes shortcomings in describing a condition predominantly managed in the outpatient setting.

The results of this work provide an insight into how our organisation delivers HCM care, and handles information, a pattern that is likely to be replicated for other hospitals delivering regional services. Our framework didn’t analyse scanned documents due to the errors associated with this document type. In the future, improved OCR techniques could allow NLP to bring content from these documents ‘into play’. We plan to integrate documents currently held outside OpenText into the pipeline so the algorithm can analyse cardiac imaging reports.

This work argues for an information management strategy that avoids scanning by using only electronic communication with external organisations, and promotes point-of-care tools with which clinicians can quickly capture diagnoses prospectively in a structured form.

### Use in Other Diseases and Institutions

Further investigation is needed to measure the algorithm’s accuracy across different clinical environments. We anticipate that the methodology’s approach would be equally useful in other diseases, especially inherited conditions due to its family-history detector component.

We recognise the need to train and validate the framework for each disease group and centre. Since the work here has dealt with most general errors, we anticipate that training requirements will fall over time. The algorithm’s performance will continue to improve as it receives knowledge from training feedback.

We will also need to address technical barriers to deployment on other sites. The pipeline is ideally performed with server infrastructure. However, its low resource requirements mean it could be run on a standard hospital computer where running time would be proportional to the performance capacity of the machine. We predict this would take around a week to run in practice.

### Limitations

Misclassification could result from transcription errors arising between clinic letters. If AF and HF, but not HCM, occurred in documents that post-date those which contain the HCM diagnosis, these complications will be missed. Patients may also be incorrectly classified. In the future, this could be mitigated by using confirmatory data from genetic or cardiac imaging results.

## Conclusions

In this study, our NLP framework improved the accuracy of patient lists by discovering more than 700 additional patients not known to any structured resource, including those managed by the specialist service. It characterised patients with two disease complications, no cardiology follow-up, and identified individuals likely being screened. This work revealed the limitations of routine hospital records and disease registries for teams moving to a PHM model of care. Although the use of NLP creates fresh challenges around information management, the potential payoff for targeted and responsive care is substantial.

## Data Availability

Software used to run the analysis is available as open source software. Vocabulary is included as appendix. Data is not available, as it describes patients.

https://github.com/reality/komenti

## Acknowledgements

We are grateful to Ameenathal Fawzy, Asgher Champsi, Rubia Akhtar, Haroon Ahmad, Noreen Akram, and Dino FA Motti for performing initial validation. We would also like to thank Andreas Karwath, Paul Schofield, and Egon Willighagen for useful discussions surrounding the NLP technology. This work uses data provided by patients and collected by the NHS as part of their care and support.

## Sources of Funding

GVG and LTS acknowledge support from the NIHR Birmingham ECMC, NIHR Birmingham SRMRC, Nanocommons H2020-EU (731032) and the NIHR Birmingham Biomedical Research Centre and the MRC HDR UK (HDRUK/CFC/01), an initiative funded by UK Research and Innovation, Department of Health and Social Care (England) and the devolved administrations, and leading medical research charities. The views expressed in this publication are those of the authors and not necessarily those of the NHS, the National Institute for Health Research, the Medical Research Council or the Department of Health.

## Appendix

### Named-entity recognition vocabularies

#### HCM

Labels: hcm, hypertrophic cardiomyopathy, cardiomyopathy, hypertrophic, hocm, hypertrophic obstructive cardiomyopathy

#### Atrial Fibrillation

Labels: atrial fibrillation, auricular fibrillation, afib, fibrillation paroxysmal atrial, fibrillation atrial, ‘fibrillations, atrial’, ‘fibrillation, auricular’, ‘fibrillations, auricular’, ‘fibrillation, atrial’, auricular fibrillations, quivering upper heart chambers resulting in irregular heartbeat, a-fib, fibrillation auriculaire, atrium; fibrillation, a fib, fibrillation atriale, fibrillation; atrial or auricular, atrial fibrillation and flutter, atrial fibrillation

#### Heart Failure

Labels: heart failure, cardiac failure, cardiac insufficiency, ‘insufficiency, cardiac’, ‘failure, heart’, failure heart, decompensation cardiac, myocardial decompensation, decompensation myocardial, insufficiency cardiac, heart insuffiency, weak heart, cardiac failures, heart; weakness, insuffisance coronarienne, cardiac function failed, ‘myocardial failure,’, insuffisance myocardique,insuffisance cardiaque, cardiac function failure, heart insufficiency, decompensation cardiaque, insuffiance cardiaque, heart weak, heart failure, myocardial failure, right ventricular failure, left ventricular failure, enlarged heart, cor pulmonale

#### Lexical Negation Vocabulary

Exclude, negative, without, no, not, deny, nothing, doesn’t, denies, denied

#### Exclusion Vocabulary

pathway, recommendations, screenings, post mortem, autopsy, date, mortem, correspondence, perspective, informed, excluding, caution in copd, risktolife, consultant, team, nurse, clinic, make sure, denies, without mention of, -ve, death, risk include, risks include, carrier, refused, rule out, negative, bloods, test, other sphingolipidosis, reduce your risk, information leaflets, 90 days of discharge, anticoagulantteam, risks involved, 999, st depression, stroke wave, stroke depression, therisksof, risk of surgery, riskofsurgery, risksofsurgery, risks of surgery, risk of, riskof, risks including, risksincluding, rule out, ruled out, adverse events, birmingham cardiomyopathy clinic, rt hcm, nhs foundation trust, should, programme, department, edgbaston, 0121, fax, specialist, nurses, non, icd check, icd checks, turtner, booklet, vaccine, information given, if you develop, heart failure model, survival score, maggic, explained that hcm, may also be considered, high risk idf, no way to prevent, target, disease/chest pain, medical centre, w33, hcm clinic, cilnic, electronically, identified the cause, impediment to surgery, grateful, services, consider referral, esc hcm risk, koine, discharge information, screening, screened, screen, screens, hcm risk, in a patient, for a patient, previously

#### Uncertainty Vocabulary

almost, suggestive, if, suggested, indicative, either, question, query, not confirmed, nothing, apparently, appeared, appearing, appears, conceivable, could, depend, depended, depending, depends, points to, check, checking, probability, unlikely, likely, review, reviewed, differential, differentials, even if, reported as normal, exclude, maybe, may be, indicate, look for, exclude, assess, excludes, arranged, suspected, screening, screened, suspicion, suspicious, distinguish, distinguishing, family studies, might, nearly, occasionally, perhaps, possible, possibly, seldom, seldomly, sometimes, somewhat, uncertain, uncertainly, can, frequently, likely, tbc, often, ought, probable, whether, probably, rarely, regularly, should, tends, usually, abeyance, abeyances, almost, alteration, alterations, ambiguities, ambiguity, ambiguous, anomalies, anomalous, anomalously, anomaly, anticipate, anticipated, anticipates, anticipating, anticipation, anticipations, apparent, apparently, appear, appeared, appearing, appears, approximate, approximated, approximately, approximates, approximating, approximation, approximations, arbitrarily, arbitrariness, arbitrary, assume, assumed, assumes, assuming, assumption, assumptions, believe, believed, believes, believing, cautious, cautiously, cautiousness, clarification, clarifications, conceivable, conceivably, conditional, conditionally, confuses, confusing, confusingly, confusion, contingencies, contingency, contingent, contingently, contingents, crossroad, crossroads, depend, depended, dependence, dependencies, dependency, depending, depends, destabilizing, deviate, deviated, deviates, deviating, deviation, deviations, differ, differed, differing, differs, doubt, doubted, doubtful, doubts, exposure, exposures, fluctuate, fluctuated, fluctuates, fluctuating, fluctuation, fluctuations, hidden, hinges, imprecise, imprecision, imprecisions, improbability, improbable, incompleteness, indefinite, indefinitely, indefiniteness, indeterminable, indeterminate, inexact, inexactness, instabilities, instability, intangible, intangibles, likelihood, maybe, may, reflect, might, nearly, nonassessable, occasionally, ordinarily, pending, perhaps, possibilities, possibility, possible, possibly, precaution, precautionary, precautions, predict, predictability, predicted, predicting, prediction, predictions, predictive, predictor, predictors, predicts, preliminarily, preliminary, presumably, presume, presumed, presumes, presuming, presumption, presumptions, probabilistic, probabilities, probability, probable, probably, random, randomize, randomized, randomizes, randomizing, randomly, randomness, reassess, reassessed, reassesses, reassessing, reassessment, reassessments, recalculate, recalculated, recalculates, recalculating, recalculation, recalculations, reconsider, consider, reconsidered, reconsidering, reconsiders, reexamination, reexamine, reexamining, reinterpret, reinterpretation, reinterpretations, reinterpreted, reinterpreting, reinterprets, revise, revised, risked, riskier, riskiest, riskiness, risking, risks, risky, roughly, rumors, seems, seldom, seldomly, sometime, sometimes, somewhat, somewhere, speculate, speculated, speculates, speculating, speculation, speculations, speculative, speculatively, sporadic, sporadically, sudden, suddenly, susceptibility, tending, tentative, tentatively, turbulence, uncertain, uncertainly, uncertainties, uncertainty, unclear, unconfirmed, undecided, undefined, undesignated, undetectable, undeterminable, undetermined, undocumented, unexpected, unexpectedly, unfamiliar, unfamiliarity, unforecasted, unforseen, unguaranteed, unhedged, unidentifiable, unidentified, unknown, unknowns, unobservable, unplanned, unpredictability, unpredictable, unpredictably, unpredicted, unproved, unproven, unquantifiable, unquantified, unreconciled, unseasonable, unseasonably, unsettled, unspecific, untested, tested, unusual, unusually, unwritten, vagaries, vague, vaguely, vagueness, vaguenesses, vaguer, vaguest, variability, variable, variables, variably, variance, variances, variant, variants, variation, variations, varied, varies, vary, varying, volatile, volatilities, volatility, not sure, investigated, investigation, investigation, investigations, monitor, monitored, development, dissociate, insufficient, future, analogous, develop, suggested, look for, imply, unless, exc, consideration, or, unkeen, discontinued

## Notes

### Competing Interest Statement

The authors have declared no competing interest.

### Author Declarations

The University of Birmingham Ethical Review granted approval (ERN_20-0338). No informed consent was required as this was a service improvement project, and the documents were not de-identified as we intend to follow up individuals lost to follow up.

## References

1. Maron BJ, Gardin JM, Flack JM, Gidding SS, Kurosaki TT, Bild DE. Prevalence of Hypertrophic Cardiomyopathy in a General Population of Young Adults: Echocardiographic Analysis of 4111 Subjects in the CARDIA Study. Circulation. 1995;92:785–789.

2. Massera D, McClelland RL, Ambale‐Venkatesh B, Gomes AS, Hundley WG, Kawel‐Boehm N, Yoneyama K, Owens DS, Garcia MJ, Sherrid MV, Kizer JR, Lima JAC, Bluemke DA. Prevalence of Unexplained Left Ventricular Hypertrophy by Cardiac Magnetic Resonance Imaging in MESA. J Am Heart Assoc Cardiovasc Cerebrovasc Dis. 2019;8.

3. Canepa M, Fumagalli C, Tini G, Vincent-Tompkins J, Day SM, Ashley EA, Mazzarotto F, Ware JS, Michels M, Jacoby D, Ho CY, Olivotto I, The SHaRe Investigators. Temporal Trend of Age at Diagnosis in Hypertrophic Cardiomyopathy: An Analysis of the International Sarcomeric Human Cardiomyopathy Registry. Circ Heart Fail [Internet]. 2020 [cited 2020 Sep 12];Available from: https://www.ahajournals.org/doi/10.1161/CIRCHEARTFAILURE.120.007230

4. Authors/Task Force members, Elliott PM, Anastasakis A, Borger MA, Borggrefe M, Cecchi F, Charron P, Hagege AA, Lafont A, Limongelli G, Mahrholdt H, McKenna WJ, Mogensen J, Nihoyannopoulos P, Nistri S, Pieper PG, Pieske B, Rapezzi C, Rutten FH, Tillmanns C, Watkins H. 2014 ESC Guidelines on diagnosis and management of hypertrophic cardiomyopathy: the Task Force for the Diagnosis and Management of Hypertrophic Cardiomyopathy of the European Society of Cardiology (ESC). Eur Heart J. 2014;35:2733–2779.

5. Ommen SR, Mital S, Burke MA, Day SM, Deswal A, Elliott P, Evanovich LL, Hung J, Joglar JA, Kantor P, Kimmelstiel C, Kittleson M, Link MS, Maron MS, Martinez MW, Miyake CY, Schaff HV, Semsarian C, Sorajja P. 2020 AHA/ACC Guideline for the Diagnosis and Treatment of Patients With Hypertrophic Cardiomyopathy. J Am Coll Cardiol. 2020;S0735109720364135.

6. Institute of Medicine (US) Committee on Assuring the Health of the Public in the 21st Century. Understanding Population Health and Its Determinants [Internet]. National Academies Press (US); 2002 [cited 2020 Nov 3]. Available from: https://www.ncbi.nlm.nih.gov/books/NBK221225/

7. Krall MA, Gundlapalli AV, Samore MH. Chapter 13 - Big Data and Population-Based Decision Support [Internet]. In: Greenes RA, editor. Clinical Decision Support (Second Edition). Oxford: Academic Press; 2014 [cited 2020 Nov 3]. p. 363–381.Available from: http://www.sciencedirect.com/science/article/pii/B9780123984760000130

8. Bean DM, Teo J, Wu H, Oliveira R, Patel R, Bendayan R, Shah AM, Dobson RJB, Scott PA. Semantic computational analysis of anticoagulation use in atrial fibrillation from real world data. PloS One. 2019;14:e0225625.

9. Shah RU, Mutharasan RK, Ahmad FS, Rosenblatt AG, Gay HC, Steinberg BA, Yandell M, Tristani-Firouzi M, Klewer J, Mukherjee R, Lloyd-Jones DM. Development of a Portable Tool to Identify Patients With Atrial Fibrillation Using Clinical Notes From the Electronic Medical Record. Circ Cardiovasc Qual Outcomes. 2020;13:e006516.

10. Weissler EH, Zhang J, Lippmann S, Rusincovitch S, Henao R, Jones WS. Use of Natural Language Processing to Improve Identification of Patients With Peripheral Artery Disease. Circ Cardiovasc Interv [Internet]. 2020 [cited 2020 Nov 2];Available from: https://www.ahajournals.org/doi/abs/10.1161/CIRCINTERVENTIONS.120.009447

11. Sammani A, Bagheri A, van der Heijden PGM, Te Riele ASJM, Baas AF, Oosters C a. J, Oberski D, Asselbergs FW. Automatic multilabel detection of ICD10 codes in Dutch cardiology discharge letters using neural networks. NPJ Digit Med. 2021;4:37.

12. Moon S, Liu S, Scott CG, Samudrala S, Abidian MM, Geske JB, Noseworthy PA, Shellum JL, Chaudhry R, Ommen SR, Nishimura RA, Liu H, Arruda-Olson AM. Automated extraction of sudden cardiac death risk factors in hypertrophic cardiomyopathy patients by natural language processing. Int J Med Inf. 2019;128:32–38.

13. Slater LT, Bradlow W, Motti DFA, Hoehndorf R, Ball S, Gkoutos GV. A fast, accurate, and generalisable heuristic-based negation detection algorithm for clinical text. Comput Biol Med. 2021;130:104216.

14. Slater LT, Bradlow W, Hoehndorf R, Motti DF, Ball S, Gkoutos GV. Komenti: A semantic text mining framework. bioRxiv. 2020;2020.08.04.233049.

15. Hansen S, McMahon M, Prat A. Transparency and Deliberation Within the FOMC: A Computational Linguistics Approach. Q J Econ. 2018;133:801–870.

16. Marstrand P, Han L, Day SM, Olivotto I, Ashley EA, Michels M, Pereira AC, Wittekind SG, Helms A, Saberi S, Jacoby D, Ware JS, Colan SD, Semsarian C, Ingles J, Lakdawala NK, Ho CY, For the SHaRe Investigators. Hypertrophic Cardiomyopathy With Left Ventricular Systolic Dysfunction: Insights From the SHaRe Registry. Circulation. 2020;141:1371–1383.

17. Ho CY, Day SM, Ashley EA, Michels M, Pereira AC, Jacoby D, Cirino AL, Fox JC, Lakdawala NK, Ware JS, Caleshu CA, Helms AS, Colan SD, Girolami F, Cecchi F, Seidman CE, Sajeev G, Signorovitch J, Green EM, Olivotto I. Genotype and Lifetime Burden of Disease in Hypertrophic Cardiomyopathy. Circulation. 2018;138:1387–1398.

18. O’Mahony C, Jichi F, Pavlou M, Monserrat L, Anastasakis A, Rapezzi C, Biagini E, Gimeno JR, Limongelli G, McKenna WJ, Omar RZ, Elliott PM, Hypertrophic Cardiomyopathy Outcomes Investigators. A novel clinical risk prediction model for sudden cardiac death in hypertrophic cardiomyopathy (HCM risk-SCD). Eur Heart J. 2014;35:2010–2020.

19. Maron MS, Rowin EJ, Wessler BS, Mooney PJ, Fatima A, Patel P, Koethe BC, Romashko M, Link MS, Maron BJ. Enhanced American College of Cardiology/American Heart Association Strategy for Prevention of Sudden Cardiac Death in High-Risk Patients With Hypertrophic Cardiomyopathy. JAMA Cardiol. 2019;4:644.

20. Neubauer S, Kolm P, Ho CY, Kwong RY, Desai MY, Dolman SF, Appelbaum E, Desvigne-Nickens P, DiMarco JP, Friedrich MG, Geller N, Harper AR, Jarolim P, Jerosch-Herold M, Kim D-Y, Maron MS, Schulz-Menger J, Piechnik SK, Thomson K, Zhang C, Watkins H, Weintraub WS, Kramer CM, Mahmod M, Jacoby D, White J, Chiribiri A, Helms A, Choudhury L, Michels M, Bradlow W, Salerno M, Heitner S, Prasad S, Mohiddin S, Swoboda P, Mahrholdt H, Bucciarelli-Ducci C, Weinsaft J, Kim H, McCann G, van Rossum A, Williamson E, Flett A, Dawson D, Mongeon FP, Olivotto I, Crean A, Owens A, Anderson L, Biagini E, Newby D, Berry C, Kim B, Larose E, Abraham T, Sherrid M, Nagueh S, Rimoldi O, Elstein E, Autore C. Distinct Subgroups in Hypertrophic Cardiomyopathy in the NHLBI HCM Registry. J Am Coll Cardiol. 2019;74:2333–2345.

21. Gorinski PJ, Wu H, Grover C, Tobin R, Talbot C, Whalley H, Sudlow C, Whiteley W, Alex B. Named Entity Recognition for Electronic Health Records: A Comparison of Rule-based and Machine Learning Approaches. ArXiv190303985 Cs [Internet]. 2019 [cited 2019 Nov 15];Available from: http://arxiv.org/abs/1903.03985

22. Chapman WW, Bridewell W, Hanbury P, Cooper GF, Buchanan BG. A simple algorithm for identifying negated findings and diseases in discharge summaries. J Biomed Inform. 2001;34:301–310.

23. Gkotsis G, Velupillai S, Oellrich A, Dean H, Liakata M, Dutta R. Don’t Let Notes Be Misunderstood: A Negation Detection Method for Assessing Risk of Suicide in Mental Health Records [Internet]. In: Proceedings of the Third Workshop on Computational Linguistics and Clinical Psychology. San Diego, CA, USA: Association for Computational Linguistics; 2016 [cited 2019 Nov 17]. p. 95–105.Available from: https://www.aclweb.org/anthology/W16-0310

